# Biological Age as a Predictor of Arterial Stiffness in Young and Early Midlife Black and White Women

**DOI:** 10.1101/2024.10.23.24316009

**Authors:** Telisa A. Spikes, Alvaro Alonso, Roland J. Thorpe, Jordan Pelkmans, Melinda Higgins, Samaah Sullivan, Sandra B. Dunbar, Charles Searles, Tené T. Lewis, Puja K. Mehta, Priscilla Pemu, Herman Taylor, Arshed Quyyumi

## Abstract

**Background:** Arterial stiffness is a precursor of hypertension and hallmark of vascular aging. Chronological age (CA), a robust determinant of arterial stiffness, is considered an insufficient indicator of aging and disparities associated with disease risk. Biological age (BA), the true age based on physiological decline, is recognized as a better indicator of disease risk however the interplay among BA, CA, and arterial stiffness remains inconclusive. We investigated the associations between BA, CA, and arterial stiffness as a potential pathway for higher cardiovascular risk among Black women and whether race modified these associations.

**Methods:** In 230 women (n=150 White, n=80 Black) enrolled in the Predictive Health Institute cohorts (age range 25-49), arterial stiffness was assessed using carotid-femoral pulse wave velocity (cfPWV), measured by applanation tonometry (SphygmoCor®). BA was estimated using the Klemera-Doubal method from 11 different clinical biomarkers and CA. Accelerated age (AccA) was calculated as the difference between BA and CA. Overall and race-specific associations between BA and arterial stiffness adjusting for sociodemographics, health behaviors, and clinical factors were estimated using multiple linear regression.

**Results:** Overall mean (SD) for CA and BA was 40 (6.1), and 38 (12.1) years, respectively. Mean (SD) for BA, AccA, and cfPWV were 43 (13.1), 1.6 (12.9), and 7.3 (1.1) in Black women and 36 (10.8), −3.2 (10.6), and 6.3 (0.8) in White women (Black-White difference in BA: 6.4 years, *P*=<0.001). Overall, BA (β=0.02 m/s per year, 95%CI: 0.00, 0.03) predicted cfPWV even after adjustment for CA, without a significant interaction by race (*P*=0.65).

**Conclusions:** BA is a predictor of arterial stiffness after accounting for CA and additional relevant factors. Although there were no racial differences in this association, both BA and arterial stiffness were higher among Black compared to White women. Longitudinal studies are needed to identify BA patterns and CVD risk in Black women.

## Introduction

Arterial stiffness, a predictive marker of future cardiovascular events, all-cause, and cardiovascular disease (CVD) mortality,^1^ is considered an important precursor of hypertension and hallmark of vascular aging.^2,3^ Vascular aging promotes remodeling of the small arteries that is associated with increased resistance, blood pressure, and ultimately central artery stiffness.^3^ Chronological age (CA) is recognized as a robust determinant of arterial stiffness,^2,4^ yet, emerging evidence suggest the sole use of CA may not be ideal for the assessment of vascular disease risk and CVD prevention.^5^ Moreover, CA represents the linear passage of time and is recognized as an insufficient indicator of aging or the disparities associated with disease risk.^6^

Biological age (BA), defined as age corresponding better to “true life expectancy” of the individual instead of CA, has been used to quantify aging-related deficits in system integrity and estimate an individual’s level of damage accrual.^7,8,9^ Despite the lack of a gold standard method to estimate BA, phenotypic measures, i.e., clinical composite biomarkers, are considered equally or more predictive of disease, disability, and mortality than molecular measures, i.e. DNA methylation, epigenetic clocks, and telomeres.^10^ Moreover, clinical composite biomarkers used to estimate BA are considered more proficient in their ability to capture systemic physiological dysregulation due to a range of factors including endogenous and exogenous stress factors, age-related molecular alterations, morbidity/mortality and health-span.^9,10^

In line with the broader evidence of race-based health disparities, racial disparities have also been found for both arterial stiffness and BA. Prior research has shown that arterial stiffness is generally higher in Black women than White women.^11,12^ Emerging findings have also shown that Black populations have higher biological ages and exhibited a faster pace of aging relative to their CA, whereas the opposite was found among White populations.^13–15^ These consequential findings may also coincide with vascular changes at the subclinical level, which has important clinical implications for both progression to clinical CVD risk and for its prevention.^16^

Given this evidence, it is plausible that higher BA may also manifest as higher arterial stiffness as a measure of vascular aging,^2^ thus contributing to the higher burden and early onset of hypertension among Black women. Investigating the interplay between BA, CA and arterial stiffness may reveal racial differences that underlie systemic changes associated with vascular aging and subclinical CVD. Thus, this study seeks to extend upon previous investigations of quantifying racial differences in aging to investigate whether aging, measured as BA, is a predictor of vascular function, measured as arterial stiffness, in a population of young adult through early midlife Black and White women free of CVD. Herein, we examined whether BA is associated with arterial stiffness and whether this association is moderated by race. We hypothesized that BA would be positively associated with arterial stiffness and that race would modify this relationship such that the slope would be greater for Black than White women.

## Methods

### Data Source and Study Design

The datasets generated during and/or analyzed during the current study are not publicly available due to ongoing data collection but are available from the senior author upon reasonable request. The Center for Health Discovery and Well Being (CHDWB) study, conducted between March 2005 to October 2009 comprised of 462 female participants aged 18 to 77 years residing in a large southeastern metropolitan area. A detailed description regarding study methods and recruitment are provided elsewhere.^17^ Briefly, the CHDWB cohort was established as an initiative aimed towards integrating predictive health principles using a combination of established and cutting-edge tools to identify and measure risks and deviations in healthy employees of two large university systems who were employed for at least two years and covered by the university sponsored health insurance plans. Participants with an acute illness, hospitalization within the past year, pregnant women, and individuals with poorly controlled medical conditions were excluded.^18^ Participants enrolled in CHDWB studies signed an informed consent that was approved by the Institutional review boards of both Emory University and Georgia Institute of Technology. All aspects of the study were approved by the Institutional Review Boards of both universities. The sample for the current cross-sectional study consisted of 230 women, aged 25-49 years at baseline. Of the 230 participants, eight women were excluded due to missing data on carotid femoral pulse wave velocity (cfPWV) (n=5) and biomarker assays (n=3) leaving a final analytic sample of 222 participants.

#### Arterial stiffness

Arterial stiffness was the outcome variable for this study and was measured as cfPWV, which is recognized as the gold standard method for the assessment of aortic stiffness ^19^. CfPWV was measured noninvasively after an overnight fast using applanation tonometry (Sphygmocor® device, Atcor Medical, Sydney, Australia) using pressure waveforms at the carotid and femoral arterial sites and calculated using standardized methods.^20^ CfPWV (meters/second) was calculated by measuring the time interval between the R-waves at each site divided by the distance. A higher PWV indicates greater arterial stiffness. Reproducibility studies in our laboratory on consecutive days demonstrated a coefficient of variation of 3.8% for cfPWV.

#### Biological Age

BA, the primary independent variable, was calculated using the Klemera-Doubal method (KDM), defined as the age at which their biological profile would be considered normal within the reference population.^7,21^ KDM BA that is greater than chronological age (CA), reflects an advanced state of biological aging, which is associated with an increased risk for disease, disability, and mortality. Conversely, a KDM BA that is less than CA is associated with a decreased risk and slower pace of aging.^21^ We calculated KDM BA based on the reformulated KDM version 2, ^21^ using 11-multisystem biomarkers that were available in our current dataset including: blood urea nitrogen, creatinine, total cholesterol, alkaline phosphatase, mean corpuscular volume, red blood cell distribution width, albumin, lymphocyte %, white blood cell count, C-reactive protein (CRP), systolic blood pressure, and CA. These biomarkers are correlated with aging and reflect the system integrity of various body systems including cardiovascular, immune, renal, hepatic, and metabolic function.^10^ We also calculated accelerated age (AccA), defined as the difference between BA and CA for descriptive purpposes. AccA reflects the pace of aging for an individual or group such that a positive value indicates that a person is biologically older than their CA and a negative value indicates a person is biologically younger than their CA for descriptive purposes. ^13,22^

The KDM BA algorithm is derived from a series of regressions of individual biomarkers on CA in a reference population. ^7,21^ The BA estimates are based upon minimizing the distance between *m* regression lines and *m* biomarker points within an *m* dimensional space of all biomarkers. ^23^ BA is computed using the KDM equation, ^7,21^ which takes information from *n* number of individual regression lines of chronological age regressed onto *n* biomarkers. Following previous work, ^14,21,22^ we formed our reference population from non-pregnant Black and White females aged 25-49 (*n*=410 Black, *n*=436 White) from National Health and Nutrition Examination Survey (NHANES) III for which data collection ran between 2017 to 2020 using the 11-biomarkers listed above.

#### Covariates

Covariates included sociodemographic, health behaviors, and clinical factors. Age, marital status (married/divorce/separated/never married/unmarried couple), self-reported race (Black/African American or White), education (≤high school, some college, or college and above), and household income (<$25K, $25K-<$50K, $50K-<$75K, & ≥$75K), were self-reported. Physical activity (none, 150 or 75 min/week of moderate-or vigorous-intensity physical activity or an equivalent combination of the two), alcohol consumption in the past 30 days (never, 1-2 times/week, 3-4 times/week, 5-6 times/week, daily, once/month, 2-3 times/month), smoking status (current/quit ≤12 months, never or quit > 12 months), history of diabetes or taking medication, and hypertension or taking medication were self-reported. Anthropometric parameters of height and weight, body mass index (BMI) kg/m^2^ calculated using the CDC guidelines, ^24^ blood pressure, and heart rate were collected by clinical staff. Psychological wellbeing, defined as depressive symptoms, was assessed using the Beck Depression Inventory Index-II (BDI-II). ^25^

#### Data Analysis

Baseline characteristics were described using mean (SD) for continuous variables, and frequency (proportion) for categorical variables. Pearson’s correlation coefficients for CA, BA, and cfPWV were calculated to determine their associations. Differences in sociodemographic, clinical and health behavior characteristics by race were evaluated using Student’s t-test for normally distributed continuous variables, Mann-Whitney U test for non-normal distributed continuous variables, and chi-square test for categorical variables. We obtained the unadjusted estimates for the association of BA with cfPWV (model 1) and used multivariable linear regression models^26^ controlling for (model 2): race, (model 3): model 2 + CA, (model 4): model 3 + other sociodemographic variables (educational attainment, income, and marital status); (model 5): model 4 + health behaviors (alcohol consumption, smoking status, physical activity); and (model 6): model 5 + clinical factors (BMI, heartrate, a comorbid variable was created and coded as 0 [no history] or 1 [history of diabetes and hypertension], and depressive symptoms). Variance inflation factors (VIFs) were conducted to check for collinearity among variables and the VIFs for all models were less than 5 indicating no presence of collinearity. Due to the inclusion of systolic blood pressure as part of the BA measure, we did not include it as a control variable for model adjustment. To determine if race modified these associations, we tested interactions between race and BA, following the same sequential adjustment as done for the primary analyses. Statistical significance was set to *p*<0.05. Calculation of BA was performed using the BioAge R package, ^21^ which allows users to parametrize measurement algorithms using custom sets of biomarkers, compare results of aging measurements to published versions of the KDM method, and to score the measurements in new datasets. All analyses were performed using STATA version 18 and R version 4.3.3. ^27^

### Results

### Sample Characteristics

Table 1 provides the distribution of participant characteristics by race. Although mean (SD) CA was similar for both groups, Black women were significantly biologically older (43.0 years, exhibited accelerated aging (1.6 years versus −3.2 years), and higher (stiffer) mean cfPWV measures (7.3 m/s versus 6.3 m/s) than White women, which were biologically younger (36.6 years). Biomarkers used to estimate BA were within normal range for both groups, but Black women had significantly higher levels of inflammation (CRP 0.26 versus 0.11 mg/dL) and lymphocyte counts (83.7% versus 70.5%), than White women. White women were more educated, had higher income, and were more likely to be married. Black women consumed less alcohol, had higher proportion of smoking, were less physically active, had higher BMI, HR, SBP, and were more likely to have been diagnosed with either diabetes or hypertension than White women.

**Table 1.**
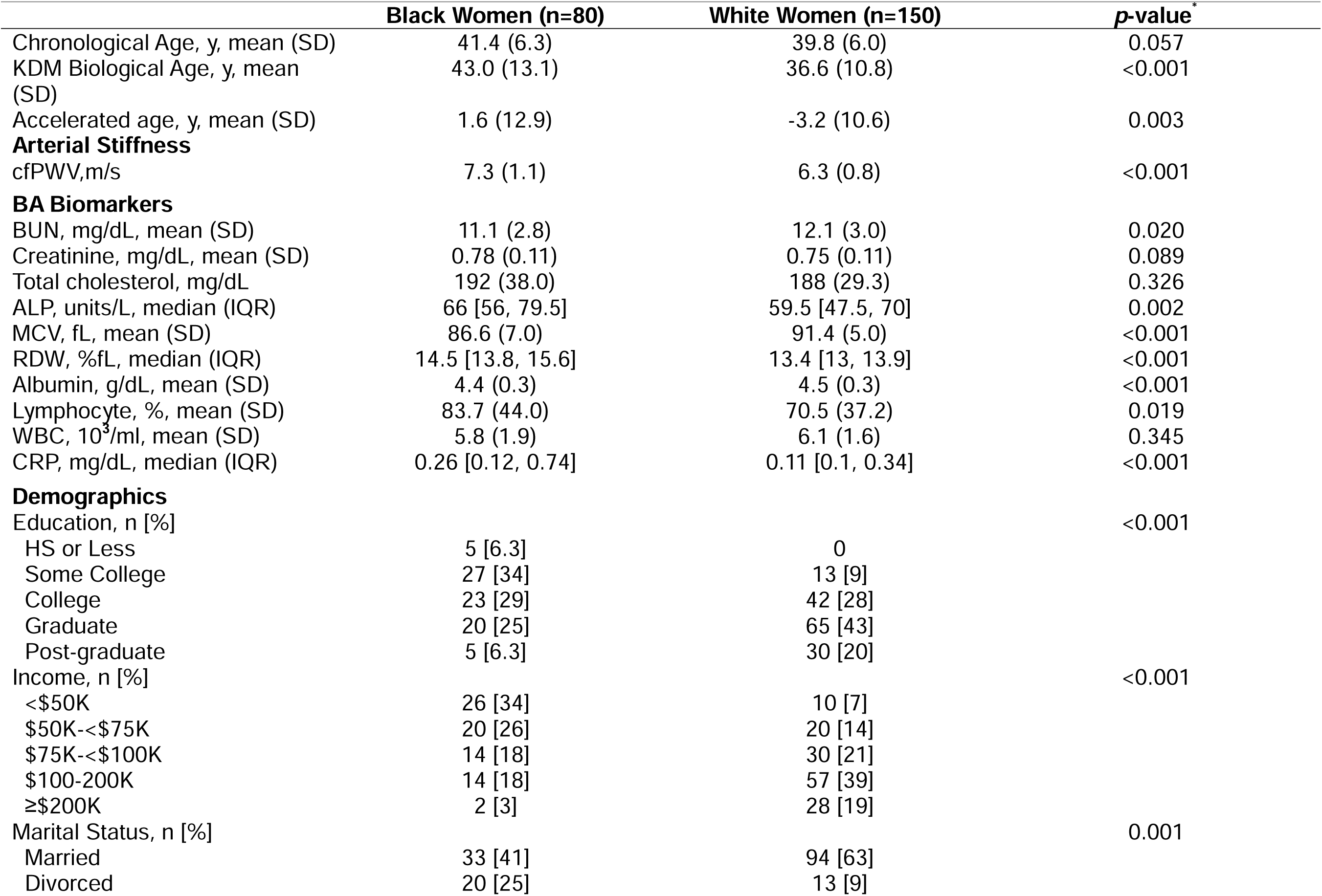
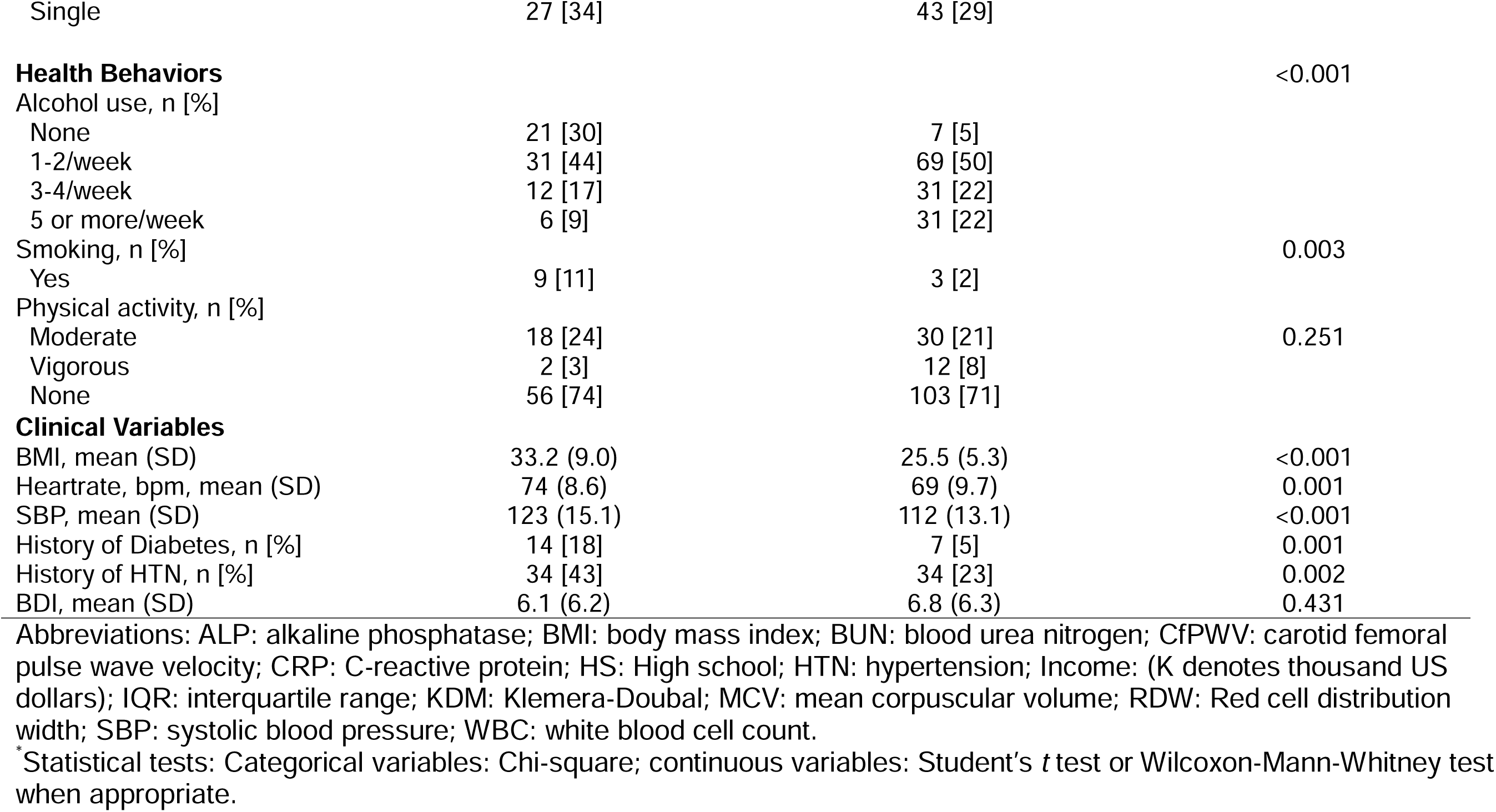
Descriptive Statistics of Participant Characteristics Stratified by Race, (n=230).

|  | **Black Women (n=80)** | **White Women (n=150)** | **p-value\*** |
| --- | --- | --- | --- |
| Chronological Age, y, mean (SD) | 41.4 (6.3) | 39.8 (6.0) | 0.057 |
| KDM Biological Age, y, mean (SD) | 43.0 (13.1) | 36.6 (10.8) | <0.001 |
| Accelerated age, y, mean (SD) | 1.6 (12.9) | -3.2 (10.6) | 0.003 |
| **Arterial Stiffness** |  |  |  |
| cfPWV,m/s | 7.3 (1.1) | 6.3 (0.8) | <0.001 |
| **BA Biomarkers** |  |  |  |
| BUN, mg/dL, mean (SD) | 11.1 (2.8) | 12.1 (3.0) | 0.020 |
| Creatinine, mg/dL, mean (SD) | 0.78 (0.11) | 0.75 (0.11) | 0.089 |
| Total cholesterol, mg/dL | 192 (38.0) | 188 (29.3) | 0.326 |
| ALP, units/L, median (IQR) | 66 [56, 79.5] | 59.5 [47.5, 70] | 0.002 |
| MCV, fL, mean (SD) | 86.6 (7.0) | 91.4 (5.0) | <0.001 |
| RDW, %fL, median (IQR) | 14.5 [13.8, 15.6] | 13.4 [13, 13.9] | <0.001 |
| Albumin, g/dL, mean (SD) | 4.4 (0.3) | 4.5 (0.3) | <0.001 |
| Lymphocyte, %, mean (SD) | 83.7 (44.0) | 70.5 (37.2) | 0.019 |
| WBC, 10<sup>3</sup>/ml, mean (SD) | 5.8 (1.9) | 6.1 (1.6) | 0.345 |
| CRP, mg/dL, median (IQR) | 0.26 [0.12, 0.74] | 0.11 [0.1, 0.34] | <0.001 |
| **Demographics** |  |  |  |
| Education, n [%] |  |  | <0.001 |
| HS or Less | 5 [6.3] | 0 |  |
| Some College | 27 [34] | 13 [9] |  |
| College | 23 [29] | 42 [28] |  |
| Graduate | 20 [25] | 65 [43] |  |
| Post-graduate | 5 [6.3] | 30 [20] |  |
| Income, n [%] |  |  | <0.001 |
| <\$50K | 26 [34] | 10 [7] | |
| \$50K-<\$75K | 20 [26] | 20 [14] | |
| \$75K-<\$100K | 14 [18] | 30 [21] | |
| \$100-200K | 14 [18] | 57 [39] | |
| ≥\$200K | 2 [3] | 28 [19] | |
| Marital Status, n [%] |  |  | 0.001 |
| Married | 33 [41] | 94 [63] |  |
| Divorced | 20 [25] | 13 [9] |  |
| Single | 27 [34] | 43 [29] |  |
| **Health Behaviors** |  |  | <0.001 |
| Alcohol use, n [%] |  |  |  |
| None | 21 [30] | 7 [5] |  |
| 1-2/week | 31 [44] | 69 [50] |  |
| 3-4/week | 12 [17] | 31 [22] |  |
| 5 or more/week | 6 [9] | 31 [22] |  |
| Smoking, n [%] |  |  | 0.003 |
| Yes | 9 [11] | 3 [2] |  |
| Physical activity, n [%] |  |  | 0.251 |
| Moderate | 18 [24] | 30 [21] |  |
| Vigorous | 2 [3] | 12 [8] |  |
| None | 56 [74] | 103 [71] |  |
| **Clinical Variables** |  |  |  |
| BMI, mean (SD) | 33.2 (9.0) | 25.5 (5.3) | <0.001 |
| Heartrate, bpm, mean (SD) | 74 (8.6) | 69 (9.7) | 0.001 |
| SBP, mean (SD) | 123 (15.1) | 112 (13.1) | <0.001 |
| History of Diabetes, n [%] | 14 [18] | 7 [5] | 0.001 |
| History of HTN, n [%] | 34 [43] | 34 [23] | 0.002 |
| BDI, mean (SD) | 6.1 (6.2) | 6.8 (6.3) | 0.431 |

Abbreviations: ALP: alkaline phosphatase; BMI: body mass index; BUN: blood urea nitrogen; CfPWV: carotid femoral pulse wave velocity; CRP: C-reactive protein; HS: High school; HTN: hypertension; Income: (K denotes thousand US dollars); IQR: interquartile range; KDM: Klemera-Doubal; MCV: mean corpuscular volume; RDW: Red cell distribution width; SBP: systolic blood pressure; WBC: white blood cell count.
\*Statistical tests: Categorical variables: Chi-square; continuous variables: Student's *t* test or Wilcoxon-Mann-Whitney test when appropriate.

#### Relationship between cfPWV and Biological Age, and Chronological Age

Pearson’s correlation coefficients between CA, BA, and cfPWV are presented in Figures 1 and 2. CfPWV was more strongly correlated with BA (r=0.36, p=<0.001) than with CA (r=0.25, p=<0.001). To determine whether BA is a predictor of arterial stiffness, we tested the association between BA and cfPWV unadjusted and adjusted for race, age, sociodemographics, health behaviors, and clinical factors. In an unadjusted model, BA was associated with a *β* =0.03 m/s per year (95% CI: 0.02, 0.04, *P*=<0.001)] higher cfPWV (Table 2). Upon adjusting for race (model 2), the association between BA and cfPWV attenuated but remained significant; *β* =0.02 m/s per year (95% CI: 0.01, 0.03, *P*=<0.001). The association between BA and cfPWV remained unchanged, *β* =0.02 m/s per year (95% CI: 0.01, 0.03, *P*=<0.001)] after adjusting for CA, Model 3, and after further adjustment for sociodemographics, health behaviors, clinical and mental health factors [model 6: *β* =0.02 m/s per year (95% CI: 0.00, 0.03, *P*=0.03)], Table 2, Models 3-6. Although race was an independent and significant predictor of cfPWV, there were no significant interactions identified between race and BA (*P*=0.65). Given the effect that diabetes and hypertension has on vascular function, we considered whether BA would be different for women diagnosed with these conditions than those without. Findings were not different than for the full cohort (data not shown).

**Figure 1.**
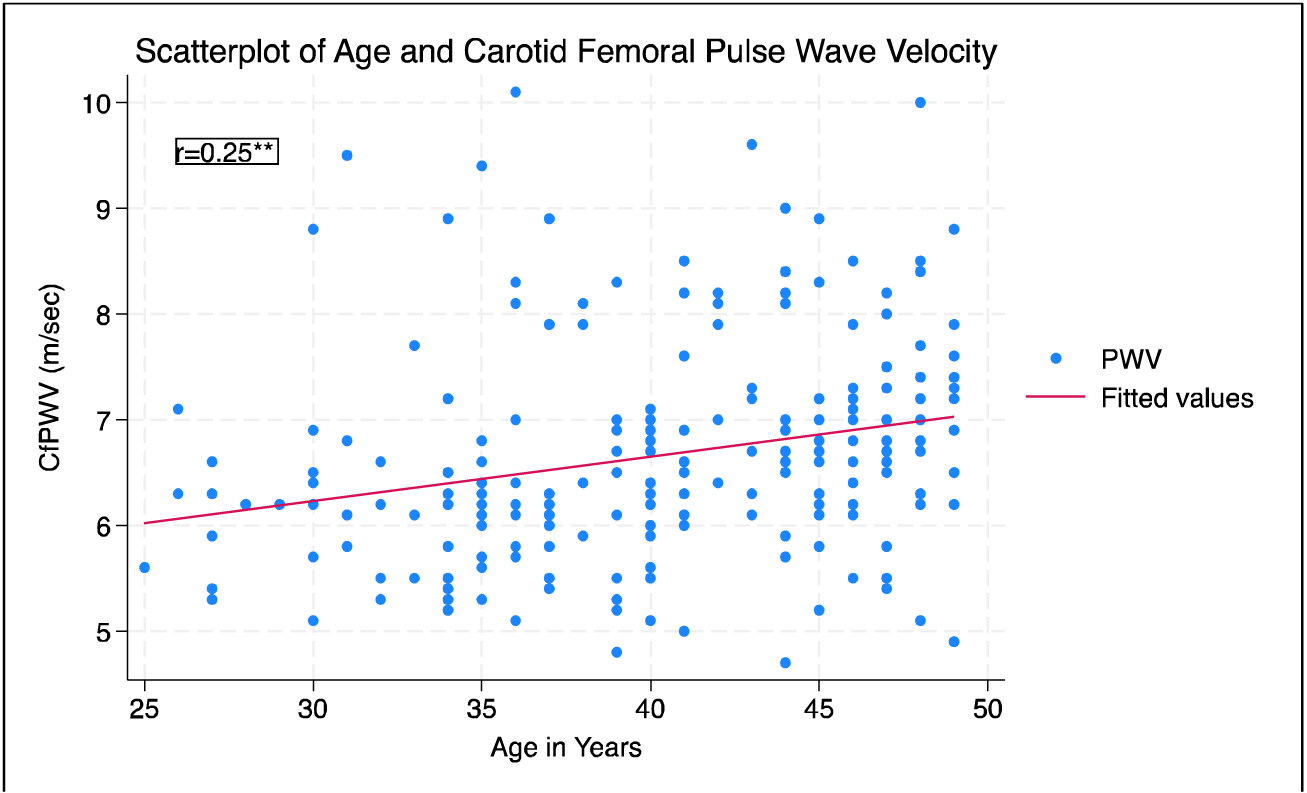
Correlations between Carotid femoral pulse wave velocity, and chronological age, (n=222). Note: **p<0.001.Abbreviations: cfPWV: Carotid femoral pulse wave velocity.

**Figure 2.**
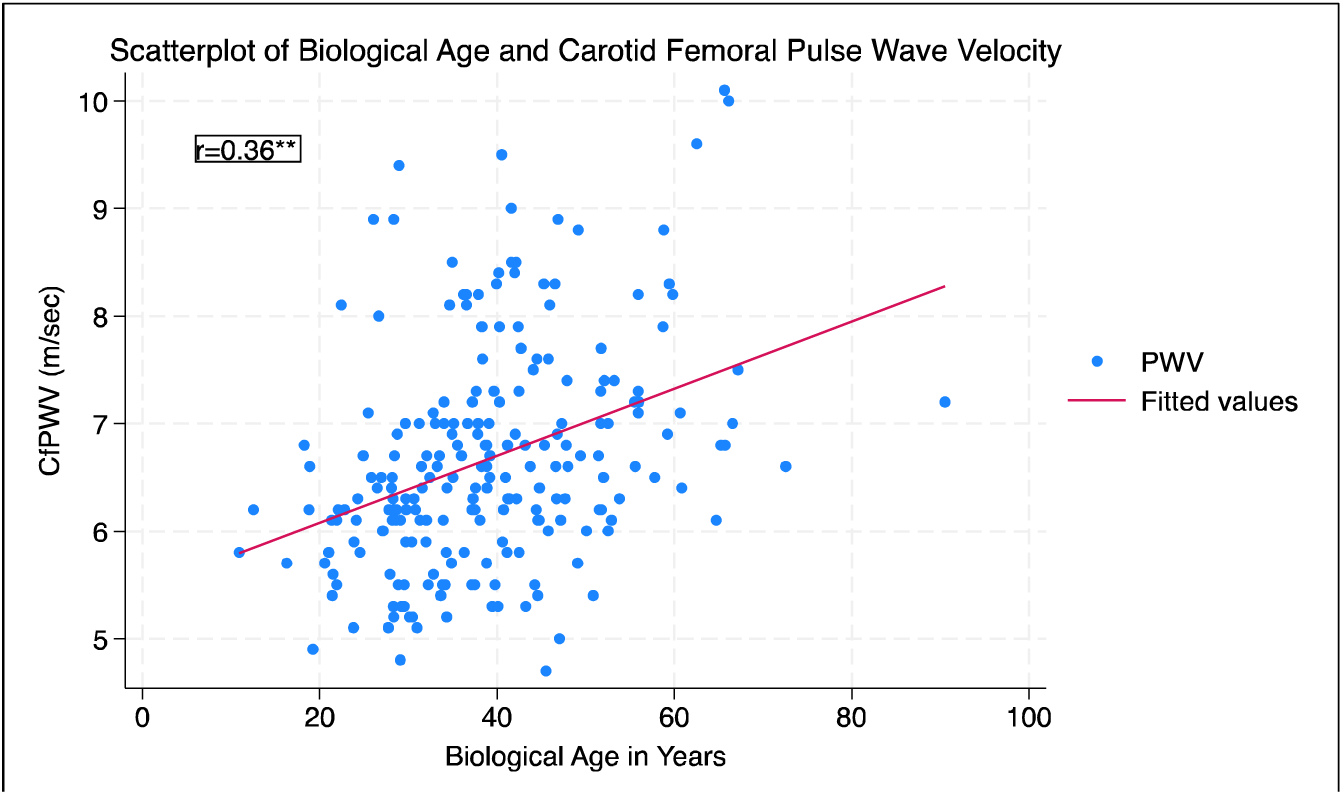
Correlations between Carotid femoral pulse wave velocity and biological age, (n=222).

**Table 2.** Association of biological age, race, and chronological age with PWV overall.

|  | Model 1 |  | Model 2 |  | Model 3 |  | Model 4 |  | Model 5 |  | Model 6 |  |
| --- | --- | --- | --- | --- | --- | --- | --- | --- | --- | --- | --- | --- |
| | $\beta$ | 95% CI | $\beta$ | 95% CI | $\beta$ | 95% CI | $\beta$ | 95% CI | $\beta$ | 95% CI | $\beta$ | 95% CI |
| Biological Age<sup>1</sup> | 0.03 | 0.02, 0.04** | 0.02 | 0.01, 0.03** | 0.02 | 0.01, 0.03** | 0.02 | 0.01, 0.03** | 0.02 | 0.01, 0.03** | 0.02 | 0.00, 0.03* |
| Race<sup>2</sup> |  |  | 0.80 | 0.54, 1.05** | 0.78 | 0.53, 1.04** | 0.63 | 0.32, 0.94** | 0.62 | 0.28, 0.97** | 0.53 | 0.16, 0.90* |
| Age<sup>3</sup> |  |  |  |  | 0.02 | 0.00, 0.04* | 0.03 | 0.01, 0.05* | 0.03 | 0.01, 0.06* | 0.04 | 0.01, 0.06* |

**[Footnote:** models included the variables listed in each model in the table]. <sup>1</sup>Klemera-Doubal Biological age. <sup>2</sup>White race reference group. <sup>3</sup>Chronological Age. Denotes $P^* < 0.05$ , $** < .0001$
Model 1: unadjusted
Model 2: model 1 + race
Model 3: model 2 + chronological age
Model 4: model 3 + sociodemographics (education-reference ≤ high school; income-reference ≤ \$25,000, marital status-reference married)
Model 5: model 4 + health behaviors (alcohol consumption, smoking status, physical activity)
Model 6: model 5 + clinical and mental health factors (BMI, heartrate, hypertension & diabetes history, and depressive symptoms).

## Discussion

In this pooled cohort of 230 Black and White women from young adulthood through middle age, we calculated BA to determine its association with arterial stiffness, measured as cfPWV, and investigated whether race, hypertension or diabetes modified these relationships. Our findings show that BA was a significant predictor of arterial stiffness among the overall cohort in both the unadjusted and fully adjusted models including race, CA, health behaviors, clinical, and mental health factors. Interestingly, despite the independent association between race and cfPWV, we did not detect any racial differences for the association between BA and cfPWV, irrespective of Black women having higher (stiffer) mean cfPWV measures than White women. With regard to the robust influence that diabetes and hypertension has on arterial stiffness, we also did not detect differences between BA among women with diabetes or hypertension from those without these conditions. Our novel findings build upon existing evidence by filling a critical gap as it relates to highlighting racial differences in BA—a risk predictive marker of overall disease-specific susceptibility and mortality ^23^—but also its synergistic relationship with arterial stiffness, a indicator of vascular aging as well as predictive marker of cardiovascular events in young adults and CVD mortality.^28,29^

Our findings that BA was a significant predictor of arterial stiffness after accounting for CA, both in the unadjusted and adjusted models for the overall cohort aligns with previous investigations that have identified associations between BA, psychosocial factors, and all-cause and CVD mortality.^13,15^ Importantly, after adjusting for other key confounders of race, age, health behaviors, and CVD risk factors, this association persisted indicating these factors may be important mediators between BA and cfPWV. The aging process is complex and estimation methods utilizing a composite of clinical biomarkers such as the KDM can provide a broader overview of the aging process that goes beyond a single body system.^30^ Additionally, BA can provide the true aging of the body system beyond the shared characteristics of CA for a given population as in our cohort. The synergistic investigation of vascular functional biomarkers such as pulse wave velocity and BA demonstrates the interconnectedness of the aging process. Moreover, the data is especially useful for the assessment and targeting for CVD prevention efforts as well as identification of critical upstream factors that may influence these disparate patterns, thereby expediting the onset of hypertension and CVD events.

Our hypothesis that race would modify the association between BA and cfPWV was not supported despite evidence showing that mean cfPWV measure was higher among Black women compared to White women. This could be due, in part, to the smaller sample of Black women in this cohort, thus limiting our ability to detect significant differences in these associations. An absence of evidence does not translate to absence of an association and thus future studies with larger sample sizes of Black women are needed to fully investigate these relationships. ^31^ Previous investigations have found racial differences in BA such that Black populations were biologically older compared to White populations,^13^ while other studies have found social hallmarks of aging^32^ including depressive symptoms and lower social class as determinants of accelerated aging in Black populations.^14^ Likewise, studies have also consistently shown that Black women have significantly stiffer arteries than White women even after accounting for CA and risk factors.^11,12,33^ Nonetheless, our findings are concerning, particularly for Black women, since these women were free of CVD at enrollment, suggesting these women have begun exhibiting evidence of vascular aging despite their younger CA.

These findings may partially underlie why Black women have some of the highest CVD associated risk factors, poor CVD outcomes, and shorter life expectancy compared to White women.^34^ According to the *Weathering Hypothesis,* the stress of living in a racially unjust society is a significant contributor of the physiological wear and tear of body systems, thus contributing to the premature onset of morbidity and mortality of racial ethnic populations.^35^ The older BA observed among Black women in the current study may be evidence of this weathering phenomenon; however, we were unable to test this hypothesis due to the lack of variables associated with social hallmarks of aging in the current dataset. Consideration of social factors that distinguish the dichotomous social context associated with race, and that are also unique among Black women, may be key in disentangling the effect of Black race as a determinant of arterial stiffness and worse CVD outcomes for Black Americans. Greater emphasis directed at providers focused on earlier CVD risk, identification, and detection efforts may collectively mitigate the incidence of premature CVD morbidity and CVD events,^36^ thus promoting healthier aging while reducing the economic burden associated with the management of CVD and other comorbid diseases.

### Clinical Implications

Arterial stiffness is considered a composite measure of what life has done to the vascular system ^3^, and as such, further study of biobehavioral mechanisms are needed to fully elucidate potential pathways that may affect equitable and healthy aging. Black women in our cohort were biologically older and exhibited a faster pace of aging than White women. The consequences of accelerated aging is postulated to result in the earlier onset of chronic disease and mortality that is commonly associated with older age. ^37^ This process may also serve as an important contributor to the ever-increasing burden of hypertension among Black women which has now outpaced that of Black men, (58.4% versus 57.5%).^38^ Furthermore, the effect of vascular aging has also been linked to the development of left ventricular diastolic dysfunction and impaired ventricular-arterial coupling among women and not men, which is also suggested to promote the development of heart failure.^39^ Thus, the inclusion of Black women in clinical research is of high priority.

#### Strengths

To our knowledge, our study is the first of its kind to investigate BA utilizing clinical biomarkers to quantify its influence on arterial stiffness in a cohort of women under 50 years of age free of CVD. Clinical markers used to estimate BA, as used in the current study, are considered more robust than epigenetic measures as they better capture physiological dysregulation due to exogenous and endogenous stress factors, age related molecular alterations, morbidity and mortality risk, and health span. ^10,21^ Moreover, clinical biomarkers are affordable and suitable for detecting signs of risk decades before disease onset as well as evaluating the efficacy of interventions.^10^ This is important given that Black women have remarkably higher rates and are more likely to acquire hypertension at an earlier age than aged-matched White women.^36,40^ The ability to examine these measures synergistically offers insight into the cardiovascular disparities that continue to disproportionately impact Black women. Additionally, our cohort included Black women, as prior studies that have focused on aging and health have been limited by a lack of racial/ethnic diversity, ^41^ thus leaving a significant gap in our knowledge regarding its influence on racial health disparities.

#### Limitations

In view of our strengths, we also had some limitations that must be acknowledged. We were unable to investigate the relationships between AccA and arterial stiffness longitudinally to identify patterns of aging over time. Assessment of BA and AccA with health outcomes yields more benefit when BA is assessed at baseline and AccA longitudinally to identify variations in aging and health patterns with respect to health outcomes. ^22^ Our sample size was small and likely underpowered to detect a difference in BA by race as well as by comorbid conditions and therefore larger sample sizes are needed to begin disentangling these differences. Unlike cfPWV, there is not a gold-standard method for estimating BA and therefore identifying the best method for risk prediction may not be suitable for use across all populations. Finally, our sample was relatively healthy from a single metropolitan area and therefore is not nationally representative of the general population.

## Conclusions

In this bi-racial population-based cohort study of early and middle-aged women, BA was associated with arterial stiffness after accounting for clinical CVD risk factors. Measurement of BA rather than CA provides a better index of vascular health in women. Although Black race was an independent determinant of arterial stiffness, race did not modify the associations between BA and arterial stiffness. Longitudinal studies are needed to compare and evaluate different aging measures and its utility as a predictor of CVD, which has important clinical implications for the prevention of CVD, especially in high-risk groups.

## Data Availability

The datasets generated during and/or analyzed during the current study are not publicly available due to ongoing data collection but are available from the senior author upon reasonable request.

## Acknowledgements

We would like to thank the participants for their contribution to this work as well as the research staff.

